# Associations of Global Country Profiles and Modifiable Risk Factors with COVID-19 Cases and Deaths

**DOI:** 10.1101/2020.06.17.20133454

**Authors:** Samuel Joseph Burden, Josefien Rademaker, Benjamin David Weedon, Luke Whaymand, Helen Dawes, Alexander Jones

**Affiliations:** Centre for Movement, Occupational and Rehabilitation Sciences, Oxford Institute of Nursing, Midwifery and Allied Health Research, Oxford Brookes University, UK; Department of Paediatrics, University of Oxford, UK; Leiden University Medical Centre, Netherlands; Oxford Health NHS Foundation Trust

## Abstract

Modifiable risk factors affect SARS-CoV-2 infection and mortality raising the possibility that lifestyle modification could play a role. This has not been studied at a global level. We analysed publicly available data from countries reporting COVID-19 cases and deaths. Associations of modifiable risk factors with total cases and excess deaths were determined with and without adjustment for confounders. 4,670,832 cases and 311,384 deaths were reported by 181 countries by 18th May 2020. Wealthier countries had the greatest caseload. Obesity was the primary modifiable risk factor for infection and greater age, male sex, physical inactivity and low salt consumption were associated with excess deaths. Obesity was less influential on mortality than physical inactivity. Globally, obesity confers vulnerability to SARS-CoV-2 infection and physical inactivity likely explains the greater mortality in the obese. High salt consumption may induce reductions in tissue ACE2 expression and subsequently reduce mortality rates.

## Introduction

In December 2019, a novel coronavirus infection (Severe Acute Respiratory Syndrome Coronavirus 2; SARS-CoV-2) broke out in Wuhan, China (1), causing a disease known as Coronavirus Disease-2019 (COVID-19). On 11^th^ March 2020, the World Health Organization (WHO) announced that the outbreak had reached pandemic status. As of the 18th May 2020, a total of 4,670,832 cases and 311,384 deaths have been reported worldwide by the European Centre for Disease Prevention and Control (ECDC) (2).

The transmission and associated outcomes of the SARS-CoV-2 virus have been studied, including the effect of comorbidities and modifiable risk factors such as older age, obesity, diabetes and high blood pressure (HBP). In the largest study to date with more than 5,600 reported COVID-19 deaths, Williamson *et al* found that such factors were associated with more COVID-19 death (3). Smaller studies report similar findings (4, 5). However, these studies are country-specific and do not address the pandemic on a global scale. Other factors, including physical inactivity (PI), have not been studied in this context, despite strong links with obesity and morbidity and mortality from traditional causes (6, 7). The known benefits of reducing PI, such as improved immune function and fewer respiratory tract infections (8), might also apply in SARS-CoV-2 infection (9). Dietary salt intake has recently been suggested as a potential factor in the pathogenesis of COVID-19 (10). The influence of salt on expression of Angiotensin-Converting Enzyme 2 (ACE2) receptors, which are essential to viral entry into host cells, offers a possible explanation for this (11, 12). Beyond modifiable risk factors, the effect of characteristics such as healthcare access and wealth have not been studied in relation to COVID-19, despite a likely influence.

International efforts have been made to ensure that COVID-19 outcome data are made publicly available. Therefore, we aimed to collate such data and establish whether modifiable risk factors and country-specific characteristics are associated with the number of COVID-19 cases and deaths. Such information might be useful in the management of the current and of future viral pandemics.

## Methods

All cross-sectional data for countries that made them available to public sources were collated. Associations of pre-identified confounders and common modifiable risk factors with the total number of COVID-19 cases and deaths for each country were tested. A smaller number of countries provided outcome data by sex, which were analysed separately to confirm that findings in the larger analysis were not biased by using combined outcome data for men and women.

### COVID-19 Cases and Deaths

Data were obtained on the 18th of May 2020 from ECDC, which provides daily totals for the number of COVID-19 cases and deaths for most countries (2). The ECDC collects data from various health authorities worldwide and 500 relevant sources are screened daily to collect the latest figures. Each data entry is then validated and published on the ECDC database, which can be freely accessed and downloaded.

Data for outcomes in men and women separately were obtained on the 20th May 2020 from Global Health 5050, which provides weekly totals for countries that report such data (13). Data from each country are collected from the last available date and may differ from the larger and more frequently updated ECDC dataset.

Daily figures were summed to give the total number of cases and deaths per country. Excess deaths were defined as the number of deaths above or below those linearly predicted by case number for the whole population. This was repeated on a sex-specific basis in the smaller dataset.

### COVID-19 Risk Factors

To determine the effect of risk factors, data on the agedness of the population and the prevalence of modifiable risk factors were obtained.

#### Agedness

The 2019 Revision of World Population Prospects is an official United Nations (UN) estimate of key demographic data that provides the total population of men and women in each 5-year age group in each country (14).

Agedness was calculated for each country as the number of individuals aged ≥ 65 years divided by the number of individuals < 65 years-old. This ratio is an internationally recognized method for determining agedness of the population and is used frequently in national statistics (15).

#### Modifiable Risk Factors

The prevalence of modifiable risk factors in each country was obtained from the World Health Organization (WHO) Noncommunicable Diseases Country Profiles 2018 (16). These data include alcohol intake, PI, salt intake, current tobacco smoking, HBP, diabetes and obesity and were collected for men and women separately in all WHO member states. Many of these risk factors coexist in individuals, as part of, for example, the metabolic syndrome, but are addressed here separately to identify the main drivers for COVID-19 cases and deaths. Their definitions can be found in Table S1.

#### Country Size and Affluence

Data on country population and land area were used to control for the effect of large populations and population density. Population was obtained from the UN 2019 Revision of World Population Prospects and land area (km^2^) was obtained from The World Bank (14, 17). Statistical adjustment for both of these parameters together controls for population density.

The affluence of a country was measured using gross domestic product per person (GDPP) and the healthcare access and quality index (HAQ). GDP per country in 2018 was obtained from The World Bank and was divided by population size to provide GDPP (18). The HAQ was obtained for each country from the Global Burden of Disease Study 2016 and is amply described elsewhere (19). Briefly, HAQ calculates an index between 0-100, based on deaths from 32 causes that should not occur in the presence of effective healthcare access and quality, with 100 being the best score.

### Statistics

Statistical analysis was performed using STATA (version 14.2, StataCorp, College Station, Texas). Data were checked for normality by visual inspection of histogram plots and log-transformed where necessary. Results are presented as median [interquartile range (IQR)] and compared across sexes using the Wilcoxon signed-rank test. Univariate linear regressions were performed to assess the effect of independent variables on the numbers of cases and excess deaths across countries (model 1). Multiple linear regressions were then performed to adjust for the effects of pre-defined confounders (agedness, GDPP, HAQ, population and land area) (model 2). Pearson correlation coefficients (r) were used to allow for comparison of the strength of associations, together with their associated P-values. P < 0.05 was considered statistically significant.

## Results

Data on cases and excess deaths were available for 181 countries. Data on confounders were available for 167-181 countries and on risk factors for 133-168 countries, depending on the variable. Sex-specific data on cases were available for 55 countries and on deaths for 47, with 33-36 countries having sufficient data for risk factor assessment in both models, depending on the variable.

A total of 4,670,832 cases and 311,384 deaths due to SARS-CoV-2 were reported by 18th May 2020. A summary of the outcome data is presented in Table 1. Across countries, there were a median(IQR) of 1,421 (285, 10,610) cases, 28 (4, 263) deaths and 1.21 (0.41, 2.24) excess deaths per case. In the sex-specific outcome dataset, case rates did not differ between men and women [6,757 (2,485, 23,204) vs. 6,338 (1,933, 17,142), P = 0.73]. However, male sex was associated with nearly twice as many deaths compared to women [481 (143, 2,970) vs. 277 (120, 1,670), P < 0.001]. As excess deaths were modelled separately for men and women in this dataset, there was no gender difference for this measure, by design.

**Table 1.** Summary Table of SARS-CoV-2 Outcome Data

|  | Overall | Men | Women | *P* * |
| --- | --- | --- | --- | --- |
| **Cases** |  |  |  |  |
| N | 181 | 55 | 55 |  |
| Median (IQR) | 1421 (285, 10610) | 6757 (2485, 23204) | 6338 (1933, 17142) | 0.73 |
| **Deaths** |  |  |  |  |
| N | 181 | 47 | 47 |  |
| Median (IQR) | 28 (4, 263) | 481 (143, 2970) | 277 (120, 1670) | < 0.001 |
| **Excess Deaths** |  |  |  |  |
| N | 181 | 37 | 37 |  |
| Median (IQR) | 1.21 (0.41, 2.24) | 1.08 (0.70, 1.74) | 1.08 (0.70, 1.59) | 0.79 |

\* Men vs. Women; N, number of countries; IQR, interquartile range.

Alcohol, salt intake, tobacco use and HBP were all significantly higher in men compared to women whereas agedness, PI and adult obesity were all significantly lower (P < 0.001 for all). There were no sex differences between men and women for diabetes (Table 2).

**Table 2.** Summary Table of Sex-Specific Independent Variables

|  | Men | Women | *P* |
| --- | --- | --- | --- |
| **Agedness** |  |  |  |
| N | 181 | 181 |  |
| Median (IQR) | 7.3 (3.2, 15.1) | 8.8 (4.0, 20.7) | < 0.001 |
| **Alcohol Use (L.person<sup>-1</sup>.Year<sup>-1</sup>)** |  |  |  |
| N | 168 | 168 |  |
| Median (IQR) | 11 (4, 16) | 2 (1, 3.5) | < 0.001 |
| **Physical Inactivity (%)** |  |  |  |
| N | 140 | 141 |  |
| Median (IQR) | 24 (16, 31) | 33 (24, 42) | < 0.001 |
| **Salt Intake (g.day<sup>-1</sup>)** |  |  |  |
| N | 167 | 167 |  |
| Mean* (IQR) | 9.3 (7, 11) | 8.4 (7, 10) | < 0.001 |
| **Tobacco Use (%)** |  |  |  |
| N | 133 | 136 |  |
| Median (IQR) | 30 (21, 41) | 6 (2, 18) | < 0.001 |
| **High Blood Pressure (%)** |  |  |  |
| N | 167 | 167 |  |
| Median (IQR) | 23 (21, 28) | 21 (18, 24) | < 0.001 |
| **Diabetes (%)** |  |  |  |
| N | 167 | 167 |  |
| Median (IQR) | 8 (6, 10) | 8 (6, 10) | 0.55 |
| **Adult Obesity (%)** |  |  |  |
| N | 167 | 167 |  |
| Median (IQR) | 16 (5, 24) | 24 (12, 29) | < 0.001 |

\* Men and women had identical medians so mean values were used to highlight the difference; IQR, interquartile range.

## COVID-19 Cases

### Sex-combined outcome data

Population, GDPP, land mass, HAQ and agedness were all positively associated with the number of COVID-19 cases in univariate analyses (Table S2, model 1). However, after mutual adjustment (model 2), only population, GDPP and HAQ remained significant (Figure 1).

**Figure 1.**
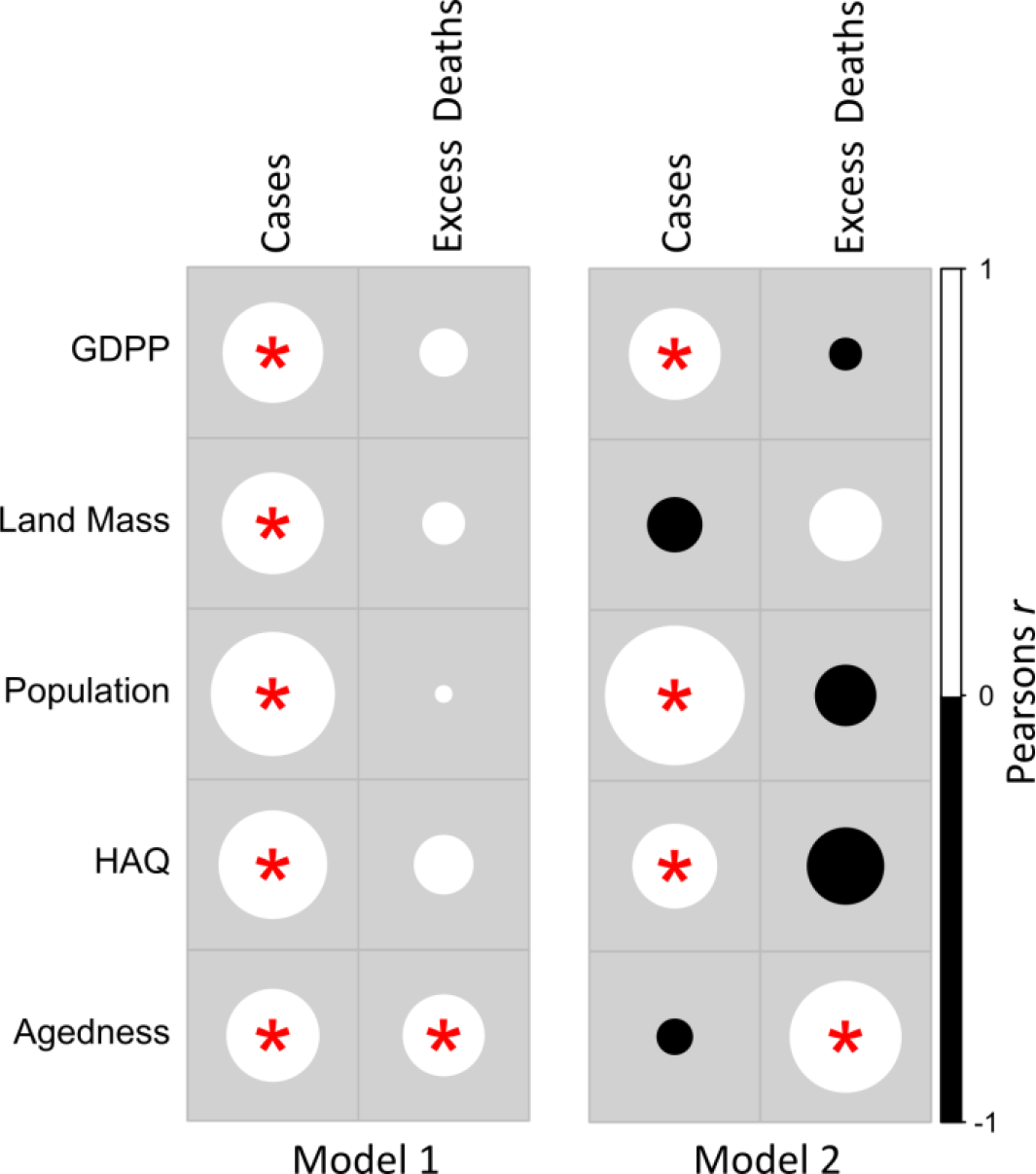
Correlations of COVID-19 Cases and Excess Deaths with Country Characteristics. Circle size represents the strength of the Pearson Correlation Coefficient with white and black representing a positive and negative correlation, respectively. Model 1 univariate analysis and Model 2 adjusted for gross domestic product per person (GDPP), population, land mass, healthcare access and quality index (HAQ) and agedness. Red star (*) indicates P<0.05.

All of the modifiable risk factors were positively related to the number of COVID-19 cases in univariate analyses, except tobacco in men, diabetes in women and HBP in either sex, where there were no significant relationships as shown in Figure 2A (Table S3, model 1). After adjustment for confounders (model 2), only obesity in men remained significantly associated with cases (Figure 2B).

**Figure 2.**
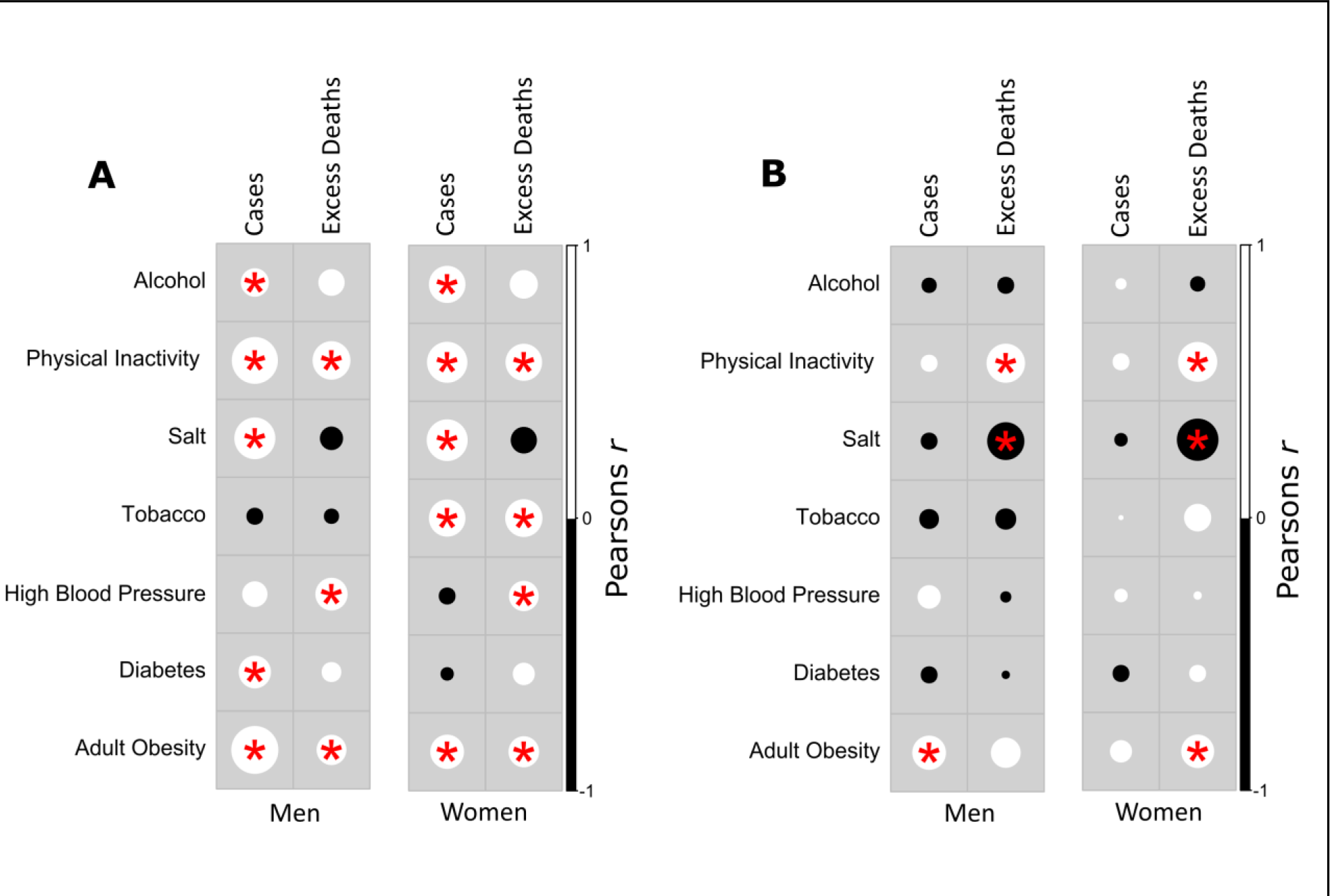
Correlations of COVID-19 Cases and Excess Deaths with Modifiable Risk Factors for Men and Women. **(A)** Model 1 univariate and **(B)** Model 2 adjusted for gross domestic product per person (GDPP), population, land mass, healthcare access and quality index (HAQ) and agedness. Circle size represents the strength of the Pearson Correlation Coefficient with white and black representing a positive and negative correlation, respectively. Red star (*) indicates P<0.05.

### Sex-specific outcome data

When associations of risk factors with the smaller dataset of sex-specific outcome data were tested (Table S4), obesity in both men and women were predictive of cases after adjustment in model 2.

Higher salt intake in men and women was also associated with fewer cases in model 2 but no other variables were significant after adjustment for confounders.

## COVID-19 Excess Deaths

### Sex-combined outcome data

Of the potential confounders, only agedness was associated with the number of excess deaths in model 1 (Table S3) and this positive association remained significant after adjustment for confounders in model 2 (Figure 1).

High salt intake was not associated with excess deaths in model 1 (although the results show a statistical trend), but when confounders were adjusted for in model 2, high salt intake was associated with fewer excess deaths for both men and women as shown in Figure 2B (Table S4). PI was associated with excess deaths in both men and women in model 1 (Figure 2A) and this remained significant after adjustment for confounders (Figure 2B).

Obesity correlated with more excess deaths in both men and women in model 1 (Figure 2A), remaining significant for obese women in model 2 (Figure 2B). However, when obesity was further adjusted for the effect of PI, the relationship with excess deaths was removed (data not shown). No other risk factors were associated with excess deaths in the fully adjusted models.

### Sex-specific outcome data

In the reduced dataset, where sex-specific outcome data were available, only obesity and tobacco were associated with outcomes but these associations disappeared in the adjusted model (Table S4, model 2).

## Discussion

In this study of 181 countries with 4,670,832 COVID-19 cases and 311,384 deaths, we showed that obesity was the only major modifiable risk factor independently associated with SARS-CoV-2 infection, with a stronger association in men. Wealthier countries had more COVID-19 cases, possibly as a result of having populations with greater global mobility. Age was predictive of excess deaths while more men than women died from COVID-19. Excess deaths were associated with PI, independent from obesity, while high salt intake was associated with lower COVID-19 death.

Multiple studies have identified several risk factors for severe COVID-19 illness or death. In a study of 4,103 COVID-19 patients, predictors of hospitalization included age > 65 years, severe obesity and a history of heart failure (4). Patients were more commonly male and frequently had comorbidities such as cardiovascular disease, diabetes and obesity, compared to non-hospitalized patients (4). In the most thorough analysis to date in >17 million UK National Health Service patients and >5,600 linked COVID-19 deaths, older age, male sex, obesity, non-white ethnicity, smoking, chronic heart disease, diabetes and other such comorbidities were associated with a higher risk of COVID-19 death (3). Deprivation was also independently identified as a major risk factor (3).

Although several studies report a high prevalence of obesity in patients in intensive care (3, 20, 21), reports on the associations of COVID-19 cases and obesity are lacking. We found a greater caseload of COVID-19 in obese men and women, although this relationship was stronger in men. Furthermore, the mortality rate of COVID-19 in men was higher, consistent with earlier reports (4).

Our study cannot define the mechanisms responsible for our epidemiological findings but helps to develop some important hypotheses for further study. In particular, the higher case rates in countries with more obesity and less salt intake could support emerging hypotheses that address the effects of SARS-CoV-2 on the renin-angiotensin system. Binding to ACE2 is recognised as an important mode of host cell entry for SARS-CoV-2, which reduces ACE2 activity as a result (12). This enzyme cleaves angiotensin II (ANGII) to angiotensin-(1-7) (22), reducing ANGII activity, and is expressed by many tissues. Adipose tissue is one of these. Increased ACE2 expression in obese individuals could create a ‘viral reservoir’ in infected individuals (23), raising their likelihood of expressing symptoms and presenting to hospital and may increase the chance of being infected by offering greater opportunity for viral binding in the lungs. In support of this, raised ACE2 expression has been demonstrated in the lungs of obese mice (24). Furthermore, genes associated with lipid metabolism in lung epithelial cells from obese mice infected with SARS-CoV-2 are upregulated, potentially facilitating completion of viral replication (24). There is also evidence that men express more ACE2 than women, potentially increasing their vulnerability to SARS-CoV-2, which is consistent with our results (25).

To date, the influence of PI on vulnerability to SARS-CoV-2 has not, to our knowledge, been assessed. We found that PI was an important risk factor for excess death, even after adjusting for obesity. Once obesity was adjusted for the highly significant effect of PI on excess deaths, obesity was no longer significant. Regular physical activity lowers mortality and incidence rates for other respiratory viruses, such as influenza (26), and increased aerobic fitness is associated with improved immune function via several mechanisms including: reduction of chronic inflammation (27); increasing the proportion of naïve T-cells (28); and greater antibody responses (29). With many governments placing tight restrictions on behaviour to limit the spread of SARS-CoV-2, the impact of these policies on PI may need to be considered in light of these findings.

Despite suggestions that HBP could be an important risk factor for COVID-19 caseload and death, we did not find it to be so after adjustment for age and other confounders. This is consistent with other recent findings (3), suggesting that the association of old age with HBP could explain these initial reports.

Our most novel finding was that countries with higher salt intake had lower mortality from COVID-19. This concords with the findings of a recent unpublished paper where elevated serum sodium on routine blood tests was associated with lower odds of being admitted to intensive care (OR = 0.949 per 1 mmol/L, 95% CI: 0.911-0.987) and lower odds of receiving mechanical ventilation (OR = 0.923 per 1 mmol/L, 95% CI: 0.879-0.966) with COVID-19 (30). Post *et al* have hypothesized that reduced sodium levels may enhance cellular damage and result in a more severe COVID-19 infection (10). A high salt diet decreases myocardial ACE2 expression in the heart and other tissues, potentially reducing the opportunity for viral damage by SARS-CoV-2 (11, 31-33). This is salient, given that myocardial damage was reported as a key mechanism of death in some cohorts (5).

Despite the large study population, the lack of sex-, age- and ethnicity-specific outcome data across all countries was a limitation that we hope will soon be addressed by bodies responsible for collating such data. We addressed this for sex-specific outcomes by repeating our analyses in the subset of countries where sex-specific data were available, with reassuringly similar results. Although some data were not contemporaneous with outcomes, they were measured within the last few years and historical trends in such data (not presented) suggest that they are unlikely to have changed significantly.

In this study of global COVID-19 cases and deaths, obesity in men and women was the primary modifiable risk factor for SARS-CoV-2 infection. Greater age, male sex, PI and a low salt intake were associated with excess COVID-19 death, after adjustment for confounders. To the best of our knowledge, we are the first to show that obesity is not a risk factor for excess death when PI is adjusted for. We also showed that HBP is not associated with excess deaths once age and other confounders are adjusted for. Finally, a high salt intake was associated with lower numbers of cases and excess deaths and we hypothesize that resultant lower levels of ACE2 in the lungs and the myocardium, amongst other tissues, could mediate these findings.

## Data Availability

The data that support the findings of this study are publicly available online. These data are available from the cited references within this manuscript.

## Acknowledgements

Alexander Jones is supported by a British Heart Foundation Intermediate Clinical Research Fellowship (FS/18/22/33479). Helen Dawes is supported by the Elizabeth Casson Trust and the NIHR Oxford Health Biomedical Research Centre. Samuel Burden is supported by the Professor Nigel Groome Studentship scheme (Oxford Brookes University). The views expressed are those of the authors and not necessarily those of the NHS, the NIHR or the Department of Health.

## Supplementary

Supplementary material can be found in the supporting document for this manuscript.

